# Use of large language models as a scalable approach to understanding public health discourse

**DOI:** 10.1101/2024.02.06.24302383

**Authors:** Laura Espinosa, Marcel Salathé

**Affiliations:** Digital Epidemiology Lab, School of Life Sciences, School of Computer and Communication Sciences, EPFL, Switzerland

## Abstract

Online public health discourse is becoming more and more important in shaping public health dynamics. Large Language Models (LLMs) offer a scalable solution for analysing the vast amounts of unstructured text found on online platforms. Here, we explore the effectiveness of Large Language Models (LLMs), including GPT models and open-source alternatives, for extracting public stances towards vaccination from social media posts. Using an expert-annotated dataset of social media posts related to vaccination, we applied various LLMs and a rule-based sentiment analysis tool to classify the stance towards vaccination. We assessed the accuracy of these methods through comparisons with expert annotations and annotations obtained through crowdsourcing. Our results demonstrate that few-shot prompting of best-in-class LLMs are the best performing methods, and that all alternatives have significant risks of substantial misclassification. The study highlights the potential of LLMs as a scalable tool for public health professionals to quickly gauge public opinion on health policies and interventions, offering an efficient alternative to traditional data analysis methods. With the continuous advancement in LLM development, the integration of these models into public health surveillance systems could substantially improve our ability to monitor and respond to changing public health attitudes.

**Authors summary:** We examined how Large Language Models (LLMs), including GPT models and open-source versions, can analyse online discussions about vaccination from social media. Using a dataset with expert-checked posts, we tested various LLMs and a sentiment analysis tool to identify public stance towards vaccination. Our findings suggest that using LLMs, and prompting them with labelled examples, is the most effective approach. The results show that LLMs are a valuable resource for public health experts to quickly understand the dynamics of public attitudes towards health policies and interventions, providing a faster and efficient option compared to traditional methods. As LLMs continue to improve, incorporating these models into digital public health monitoring could greatly improve how we observe and react to dynamics in public health discussions.

## 1. Introduction

Public health is “the art and science of preventing disease, prolonging life and promoting health through the organized efforts of society” [1]. The overall aim of public health is to sustainably promote the health and wellbeing of the entire population, strengthening public health services and reducing inequalities [2], including surveillance, prevention and control of infectious diseases, promoting healthy lifestyles, researching disease, and preventing injury. Designated populations can vary, depending on context, from small neighbourhoods to global regions [3]. Moreover, some populations may be more vulnerable and at higher risk of disease due to pre-existing chronic conditions, age and developmental stage, gender and sexual identities, having disabilities or being in marginalised groups in the community (e.g., homeless people) [4].

One of the greatest achievements of public health in the 20^th^ century is vaccination against infectious diseases, which saves lives, improves individual and population health, and reduces health care costs by billions of dollars annually. Despite the success of vaccines, uptake of vaccination is still inadequate, often delayed, and unstable [5]. Understanding the factors that affect vaccine acceptance or refusal is key to increasing vaccination uptake. In October 2014, World Health Organization (WHO) Strategic Advisory Group of Experts on Immunization (SAGE) published a report of the SAGE Working Group on Vaccine Hesitancy outlying several factors that can influence vaccine acceptance or refusal grouped in four categories: populations, subgroups, communities, and individuals [6].

Social listening is “an active process of attending to, observing, interpreting, and responding to a variety of stimuli through mediated, electronic, and social channels” [7]. It helps public health experts to better engage with the population [8]. Social media has proven to be useful in public health, for example, for understanding the public perception of an epidemic [9,10]. In the context of COVID-19 pandemic, a scoping review analysed 81 studies on social media from 2020; from these and using a modified version of the framework Social Media and Public Health Epidemic and Response (SPHERE) [11], 48 studies were categorised in the public attitudes theme [12]. A systematic review performed in 2019 on existing opinion mining approaches using data from social media platforms found that 94% of the 461 included studies use data only from one social media platform and the vast majority of the studies used data from X (formerly known as Twitter), followed by Sina Weibo and Facebook [13]. Furthermore, in another systematic review of techniques for stance detection published in 2023 [14], social media was the data source of 72% of the 96 included articles in the study.

Several studies have used natural language processing algorithms to extract the general sentiment of a text [15–18], and machine learning approaches with labelled data in order to infer public health opinion from social media data. In addition, there are open-source tools available to support such processes, such as Crowdbreaks [19] and epitweetr [20] which use machine learning methods and labelled data to monitor X for extraction of stance towards public health interventions and early detection of public health threats. However, large annotated datasets are required to have enough representation of the specific tasks for ensuring proper learning and to have enough domain-specific examples when applying machine or deep learning techniques to extract or infer the stance from a text, especially in unstructured data such as social media [21]. Furthermore, having non-experts annotators will require a higher number of annotators to equal the results from the expert annotators, and increase costs [22]. These aspects may be underestimated or not properly considered when conceptualising such studies.

In recent years, the emergence of Large Language Models (LLMs) has revolutionised the field of natural language (NLP) processing. The automated identification of a text author’s position towards a specific topic (stance detection) is key in public health and other fields. LLMs, thanks to their vast training data and enormous model size with often billions of parameters, have demonstrated remarkable proficiency in this domain. A study from Brown et al., 2020, illustrated the effectiveness of models like GPT-3 in accurately determining stance from text, outperforming NLP models in both precision and recall [23]. Furthermore, the adaptability of LLMs to diverse datasets and their capacity to understand nuanced expressions have made them invaluable tools for researchers and practitioners alike. For instance, Devlin et al., 2018, highlighted the versatility of the Bidirectional Encoder Representations from Transformers in handling context-dependent stance detection tasks, setting a precedent for the more advanced LLMs [24].

More recently, LLMs have entered the public debate with the launch of ChatGPT, a chatbot originally based on the GPT-3.5 model, and more recently the GPT-4 model. These LLMs by OpenAI have triggered unprecedented public discourse around artificial intelligence across all domains. Shortly after the launch of ChatGPT, the first open LLMs were released, notably the Llama models by Meta (e.g. Llama 2 [25]). During the years of 2023 and 2024, new incumbents such as Mistral AI and Llama 3 released a number of highly performant open LLMs as a free alternative to commercially available models such as the GPT models. Early results have shown the incredible power in text understanding across domains that these models provided out of the box, pre-trained. This opened up the opportunity for the use of such models for large-scale social media analysis in many domains, including public health, without the resource-intensive steps of data annotation and model training for every individual use case.

The objective of this study was to assess how these new methods would perform for extracting the public’s stance towards vaccination using social media posts. The theme of vaccination was chosen for its relevance to public health, as well for its importance in the public discourse and the broad range of views therein. Further, our focus on stance, rather than just sentiment, is based on the notion that stance is a better measure of vaccination intention. However, stance can be more difficult to assess, because stance can be very different from sentiment (for example, the sentence “I am angry that I haven’t been vaccinated yet” expresses a negative sentiment but a positive stance towards vaccination). Finally, our choice of methods was based on practical utility and methods broadly used (or usable in the future) in various domains: annotation by experts, crowds, lexicon- and rule-based sentiment analysis tools, and large language models (LLMs).

## 2. Materials and methods

### 2.1. Data collection

Crowdbreaks was a platform that collected and filtered Twitter data (which was renamed X). Crowdbreaks used crowdsourcing annotation and trained machine learning classifiers to determine a tweet’s stance towards a specific topic (e.g., vaccination) [19].

We sampled 1,000 random English-language tweets on vaccination from tweets that Crowdbreaks collected between 2nd of December, 2019 and 11th of March, 2022. We used the following keywords: vaccine, vaccination, vaxxer, vaxxed, vaccinated, vaccinating, vaccine, overvaccinate, undervaccinate, and unvaccinated.

We used six methods to classify the user’s stance towards vaccination from those 1,000 tweets: experts, crowdsourcing, and the four large language models GPT (Generative Pre-training Transformer, versions 3.5 and 4) [26,27], Mistral 7B Dense Transformer (Mistral (7B)) [28] and Mixtral (8×7B) [29]. In addition, we used a lexicon and rule-based feeling analysis instrument named Valence Aware Dictionary and sEntiment Reasoner (VADER) [30] to classify the general sentiment from the same 1,000 tweets as an automated, non-LLM baseline.

### 2.2. Classifying the vaccine stance using experts

#### 2.2.1. Participants

We asked a convenience sample of four experts from the EPFL Digital Epidemiology Lab to identify the general sentiment and stance towards vaccination of Tweets: two public health experts (co-authors of this publication), one expert annotator, and one expert in administration. These experts were selected based on their knowledge in the field (public health), on their knowledge in the technique (annotation) and as a comparison to the crowdsourcing (expert in administration) with whom we could discuss in case of disagreements.

#### 2.2.2. Data collection

Each of the four experts individually classified the tweets according to the general sentiment and the stance of the user towards vaccination based solely on the text of the same list of tweets.

Each expert classified the tweets on a tab of a Google sheet (one tab for each expert) containing three variables: tweet text, general sentiment of the tweet, and stance towards vaccination of the tweet. Annotators were instructed not to check the other sheets of the Google file and annotated the tweets around the same time frame. The file was saved in a restricted folder in which only the selected experts could access it.

Each expert classified independently the tweets as either negative, neutral, or positive regarding the general sentiment and stance towards vaccination.

#### 2.2.3. Data analysis

We used R 4.1.3 and RStudio version 2023.06.1 for analysing the individual classifications of all tweets [31]. A new dataset was created with the following variables: tweet text, classification of the general sentiment and stance toward vaccination by each expert, agreement (binary variable with 1, if more than three experts labelled equally the tweet for each category, or 0 if it was otherwise), percentage of each class (negative, neutral, and positive) per class (general sentiment and stance towards vaccination), and agreed class (in case of disagreement, we selected the class “neutral”).

Moreover, the tweets with no partial or full agreement were labelled in two categories: the 2-2 agreement, in which two experts selected one class and two experts selected another class, and the 2-1-1 agreement, in which two experts selected the same class and the other two experts selected the remaining classes. For the 2-2 agreement, we randomly selected one of the two classes with a 50% probability and considered this the gold standard comparison. For the 2-1-1 agreement, we only performed a descriptive analysis due to the small sample size.

We analysed the proportion of each class per expert.

### 2.3. Classifying the vaccine stance using crowdsourcing

#### 2.3.1. Participants

We used crowd annotators from the platform Amazon Mechanical Turk (Mturk) [32] to classify the same list of tweets according to the stance towards vaccination. Only crowd annotators with at least 5,000 previous approved tasks and at least 98% of approval rate in the platform had access to the task. In addition, black-listed crowd annotators on the platform were excluded.

#### 2.3.2. Data collection

We published the job in Amazon Mechanical Turk (Mturk) with several information for the crowd annotators. We included the description of the task, expected time to complete the task, a question to classify the tweet based on the “attitude of the author regarding vaccines” (i.e. stance), two examples for each of the classes (positive, neutral and negative) and the indication to classify a tweet as “neutral” if the tweet was news, factual, objective or generally ambiguous.

Each crowd annotator could decide how many and which tweets to classify. Each crowd annotator could classify a maximum of 1,000 tweets, had to wait at least 2 seconds before providing the class to avoid automatic selection by a machine, and was rewarded with 0.04 USD per tweet. At the end, each tweet was classified by at least three crowd annotators.

#### 2.3.3. Data analysis

For each tweet, we had the following variables: identifier of the specific task in Amazon Mturk, tweet identifier, date of the tweet, crowd annotator identifier, tweet text, and total duration and date of the classification per tweet.

### 2.4. Classifying the vaccine stance using LLMs

#### 2.4.1. Software

We used Jupyter Notebook 6.5.2 and Python 3.9.12 to classify the same tweets according to the stance towards vaccination using GPT versions 3.5 (GPT-3.5) and 4 (GPT-4), and Ollama 0.1.17 and Python 3.11.6 to classify the same tweets according to the stance towards vaccination using Mistral (7B), Mixtral (8×7B), Llama3 (8B) and Llama3 (70B). The specific scripts are available in the repository [31]. These LLMs were selected based on its popularity of use (GPT-3.5), expected good performance (GPT-4), and usability with low computing power needed and no cost (Mistral (7B) and Llama3 (8B)) and expected good performance (Mixtral (8×7B) and Llama3 (70B)).

#### 2.4.2. Prompt engineering

We used different prompts to classify the tweets based on instructions and examples provided [S1 Prompts used for large language models, table 1]. Prompts 1 and 2 were zero-shot prompts, i.e. without labelled examples, and thus the simplest instructions provided to these LLM asking for the sentiment regarding vaccination. The remaining prompts 3 to 8 used few-shot classification, i.e., few labelled examples for each class were provided for the model to extract the features for each class and use these learned features to predict the class of new tweets [23,33,34]. Prompt 3 contained the same examples provided to crowd annotators (i.e., two examples per class). Prompts 4 to 7 were built based on the previous prompts using a random sample of 100 tweets. Prompts were tested in this sample and the wrongly classified tweets were used as examples for the following prompt. For example, if a tweet considered by experts to have a negative class was classified otherwise, this tweet was included in the next prompt as an example for the negative class. Furthermore, examples of tweets misclassified as neutral were added as exceptions for this class. All prompts except from prompt 1 requested the LLM model to provide an explanation on the classification.

**Table 1.** Main features of the eight prompts.

| Prompt | Contains examples | Number of positive examples | Number of negative examples | Number of neutral examples | Number of non-neutral exceptions | Number of general positive scenarios | Additional information |
| --- | --- | --- | --- | --- | --- | --- | --- |
| 1 | No | 0 | 0 | 0 | 0 | 0 | Nothing |
| 2 | No | 0 | 0 | 0 | 0 | 0 | Provide explanations |
| 3 | Yes | 2 | 2 | 2 | 0 | 0 | Provide explanations |
| 4 | Yes | 3 | 2 | 2 | 6 | 0 | Provide explanations |
| 5 | Yes | 8 | 2 | 2 | 6 | 3 | Provide explanations |
| 6 | Yes | 8 | 2 | 2 | 6 | 2 | Provide explanations |
| 7 | Yes | 10 | 7 | 2 | 6 | 2 | Provide explanations |
| 8 | Yes | 8 | 2 | 2 | 6 | 3 | Provide explanations and answers in specific format. Clarifications between general sentiment in tweet and stance towards vaccination |

The same parameters were used for all prompts within each LLM. For GPT-3.5, we used the following parameters: text-davinci-003 as model, 0.8 as temperature, and 0 as frequency and presence penalty. For GPT-4, we used the following parameters: gpt-4 as model, “user” as role, 0.8 as temperature, and 0 as frequency and presence penalty. For Mistral (7B), Mixtral (8×7B), Llama3 (8B) and Llama3 (70B), we used 0.8 as temperature.

### 2.5. Classifying the general sentiment using natural language processing algorithms

#### 2.5.1. Software and data collection

We used Jupyter Notebook 6.5.2 and Python 3.9.12 to classify the same tweets according to the general sentiment expressed in the tweet using VADER.

VADER assigned to each tweet a probability for having a negative, neutral and positive sentiment in the text and an overall score between −1 (most negative) and +1 (most positive).

#### 2.5.3. Data analysis

We used R 4.1.3 and RStudio version 2023.06.1 for classifying the tweets into negative, positive or neutral according to the overall score provided by VADER. Tweets with scores between −0.5 and 0.5 were classified as neutral, more than 0.5 as positive and less than −0.5 as negative. analysing the individual classifications of all tweets

### 2.6. Comparison of the classification methods

#### 2.6.1. Statistical analysis

We used R 4.1.3 and RStudio version 2023.06.1 to compare the classes for the same list of tweets provided by experts, crowdsourcing, LLMs and VADER. Experts were considered the gold standard for the comparison.

We did two comparisons based on the level of agreement by the experts: partial and full agreement in stance towards vaccination. Tweets with at least three out of four experts assigning the same class were considered to have a partial agreement. In addition, we did a comparison for each expert for all the tweets.

We calculated a confusion matrix with three classes (negative, neutral, and positive) for each pair of comparisons: experts and crowdsourcing, experts and GPT, experts and Mistral (7B), experts and Mixtral (8×7B), and experts and VADER. Moreover, a confusion matrix with three classes was also calculated for the comparison of experts and selecting the majority class (i.e., neutral) for all tweets which was considered the baseline. We calculated the overall accuracy with a 95% confidence interval and a one-side test to see if the accuracy was better than the fourth confusion matrix considering that the system would always select that majority class in the classification process.

Furthermore, for each class we calculated the sensitivity, specificity, and F1 score. For this, we used the experts classes as the gold standard and the crowdsourcing and LLM as the comparison method separately. Moreover, the baseline classifying all tweets as neutral was also applied here.

The tweets with no partial or full agreement were analysed separately.

### 2.7. Ethics

This work has been approved by the EPFL Human Research Ethics Committee (HREC No. 012.2018 / 05.03.2018).

Crowdbreaks anonymises all tweets’ text changing the mention or reference to usernames to “@username” and the URLs to “url”, and complies with the X terms of service.

We have implemented mitigation measures to ensure an ethical use of the Amazon Mturk platform and avoid any abuse of the system. We provided an overestimation of the time required for the task, provide a communication channel via email in case crowd annotators wanted to contact us, had a prompt communication in case of being contacted, accepting all jobs in maximum one day, providing comprehensive and clear description of the research context and instructions and job description, providing a monetary compensation substantially higher than the US federal government minimum wage in which the platform is based. In addition, quality criteria for selecting the crowd annotators was established to minimise the risk of having inappropriate users or users misusing the platform.

## Results

### Extracting the vaccine stance using experts

Seventy-six percent of the 1,000 tweets had a partial agreement between experts (i.e., three out of four experts agreed on the class per tweet). From these 756 tweets, 74.1%, 19.4%, and 6.5% were classified as neutral, positive, and negative stance towards vaccination, respectively.

Thirty-seven percent of those 756 tweets had a full agreement between experts (i.e., all experts agreed on the class per tweet). From these, 77.3%, 17.6%, and 5.0% were classified as neutral, positive, and negative stance towards vaccination, respectively.

Twenty-four percent of the 1,000 tweets had no partial or full agreement. From those 243 tweets, 214 had 2-2 agreement with the majority (73.8%) being labelled as neutral by two of the experts and positive by the other two experts. From the 29 tweets with 2-1-1 agreement, 10 tweets had a predominant negative or neutral stance each, and 9 had a predominant positive stance.

When analysing the individual annotations, there were differences in the distribution of the three classes. The expert annotator and one of the public health experts classified a higher number of tweets as neutral stance towards vaccination (79.6% and 87.2%, respectively), whereas the other public health expert and the expert in administration had a lower number of tweets classified as neutral towards vaccination (45.8% and 36.4%, respectively).

### Extracting the vaccine stance using crowdsourcing

A total of 467 crowd annotators classified the tweets with the majority of tweets (64.5%) being classified by three different crowd annotators, followed by 14.7% and 7.4% of the tweets being classified by 10 and four crowd annotators, respectively.

The majority of the 756 tweets had an agreement: 85.6% had a partial agreement (i.e., 75% of the crowd annotators agreed on the stance per tweet) and, from these, 91.2% had full agreement.

The stance towards vaccination of tweets with partial agreement was distributed as follows: 89.5% as neutral and 10.5% as positive, with no tweet classified as negative. The stance towards vaccination of tweets with full agreement were 98.1% classified as neutral and 1.9% classified as positive, with no tweet classified as negative (Fig 1).

**Fig 1.**
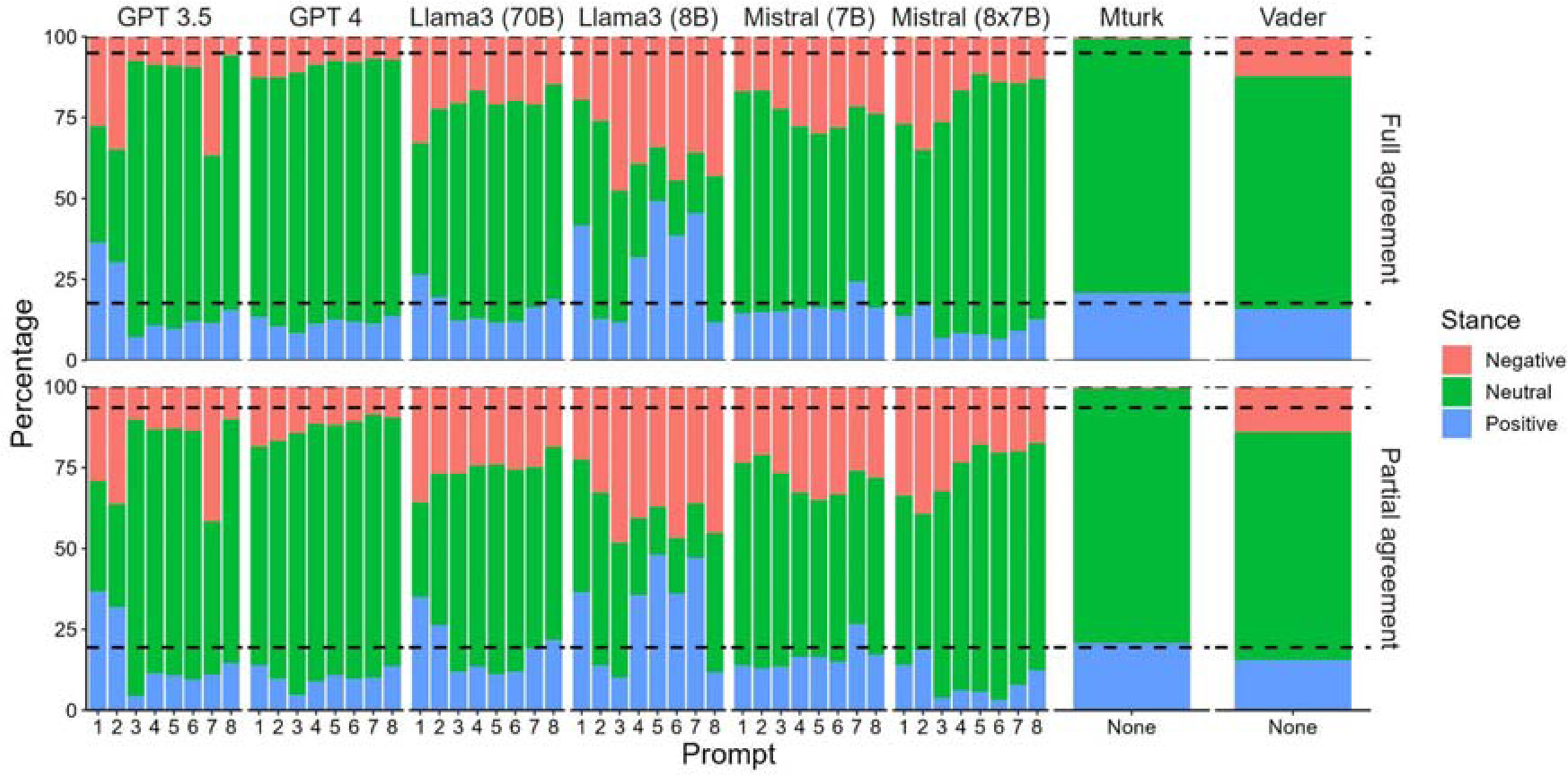
Distribution of the tweets per stance, prompt and large language model (percentage of total number of tweets). The dashed lines represent the stance distribution of positive, neutral and negative tweets according to the experts’ full and partial agreement.

### Extracting the vaccine stance using LLMs

The majority class was neutral for all prompts, LLMs and tweets with partial or full agreement by experts, except for prompts 1 of GPT-3.5, and prompts 5 and 7 of Llama3 (8B) for which the majority class was positive, and prompt 2 of GPT-3.5, prompt 1 of Llama3 (70B) and prompts 3, 4, 6 and 8 of Llama3 (8B) for which the majority class was negative (Fig 1).

### Extracting the general sentiment using VADER

The majority class was neutral (Fig 1) with an overall score ranging from −0.97 to 0.99, median of 0 and interquartile range of −0.30 and 0.01.

### Comparison of the extraction methods

#### Overall performance for tweets with partial and full agreement

The overall accuracy for all methods was worse than just selecting the majority class for all tweets, except for prompts 2-8 of GPT-4 in tweets with partial agreement among experts and for all prompts of GPT-4, prompt 3, 4 and 6 of GPT-3.5, prompts 4, 6 and 8 of Llama3 (70B) and prompts 5, 7 and 8 of Mixtral (8×7B) in tweets with full agreement among experts (Fig 2).

**Fig 2.**
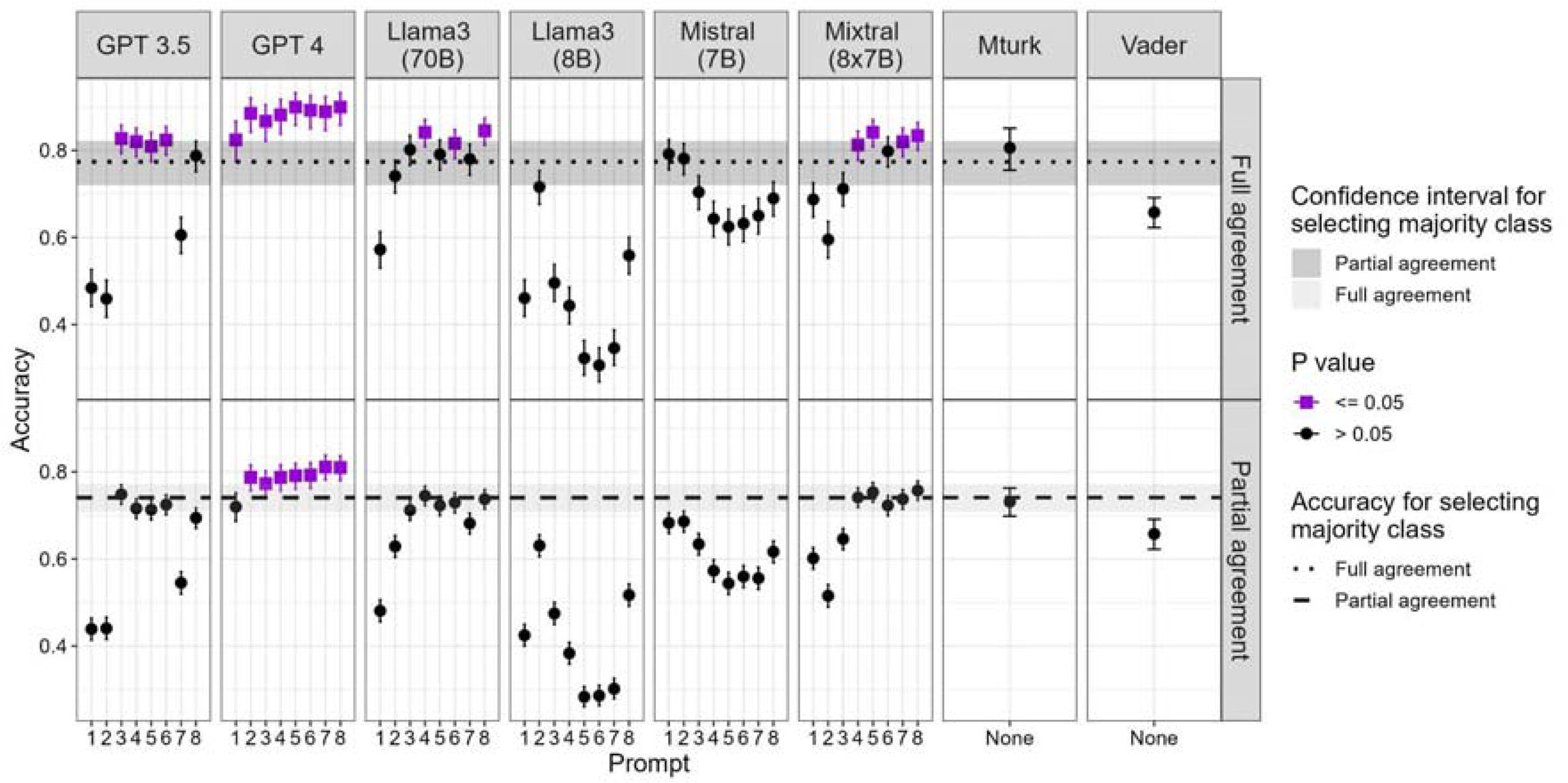
Accuracy and 95% confidence interval per prompt, model and experts’ agreement.

All prompts of GPT-4 except for prompt 1, half of the Mixtral (8×7B) prompts (4, 5, 7 and 8), prompt 3 of GPT-3.5 and prompts 4 and 8 of Llama3 (70B) performed better than crowd annotators when comparing the results with the experts’ classification in tweets with partial agreement among experts. In addition to those, prompt 1 of GPT-4, prompts 4, 5 and 6 of GPT-3.5 and prompt 6 of Llama3 (70B) performed better than crowd annotators in tweets with full agreement among experts.

Most of the prompts of Llama3 (8B), some of the prompts of GPT-3.5, Mistral (7B) and Mixtral (8×7B), and one prompt of Llama3 (70B) performed worse than extracting the general sentiment of the tweet using VADER.

Overall, Mistral (7B)’s performance was lower than that of crowd annotators. Interestingly, prompts 1 and 2 had the best accuracy with Mistral (7B) in tweets with partial and full agreement among experts, differently to the other models. Likewise, prompt 2 of Llama3 (8B) had the best accuracy among the 8 prompts.

##### Overall performance per expert

The accuracy for each prompt and model varied among experts. The experts who labelled a higher proportion of tweets as neutral stance towards vaccination had higher variability in the accuracy of each prompt within each LLM; whereas the experts who labelled a lower proportion of tweets as neutral stance towards vaccination had a more stable accuracy among the prompts within each LLM. The prompts of some LLM of only two of the experts were statistically different than just classifying all tweets neutral.

##### Overall performance for the 2-2 agreement tweets

The accuracy ranged from 0.22 (prompt 6 of Llama3 (8B)) to 0.52 (prompt 4 of GPT-4) with a similar relative difference of accuracy among prompts and methods as shown for the full- or partial-agreement tweets.

#### Performance per class for partial and full agreement

The performance metrics per class and method showed high variability according to the experts’ agreement on stance towards vaccination with an overall highest performance for neutral tweets based on the F1 score and sensitivity (Fig 3).

**Fig 3.**
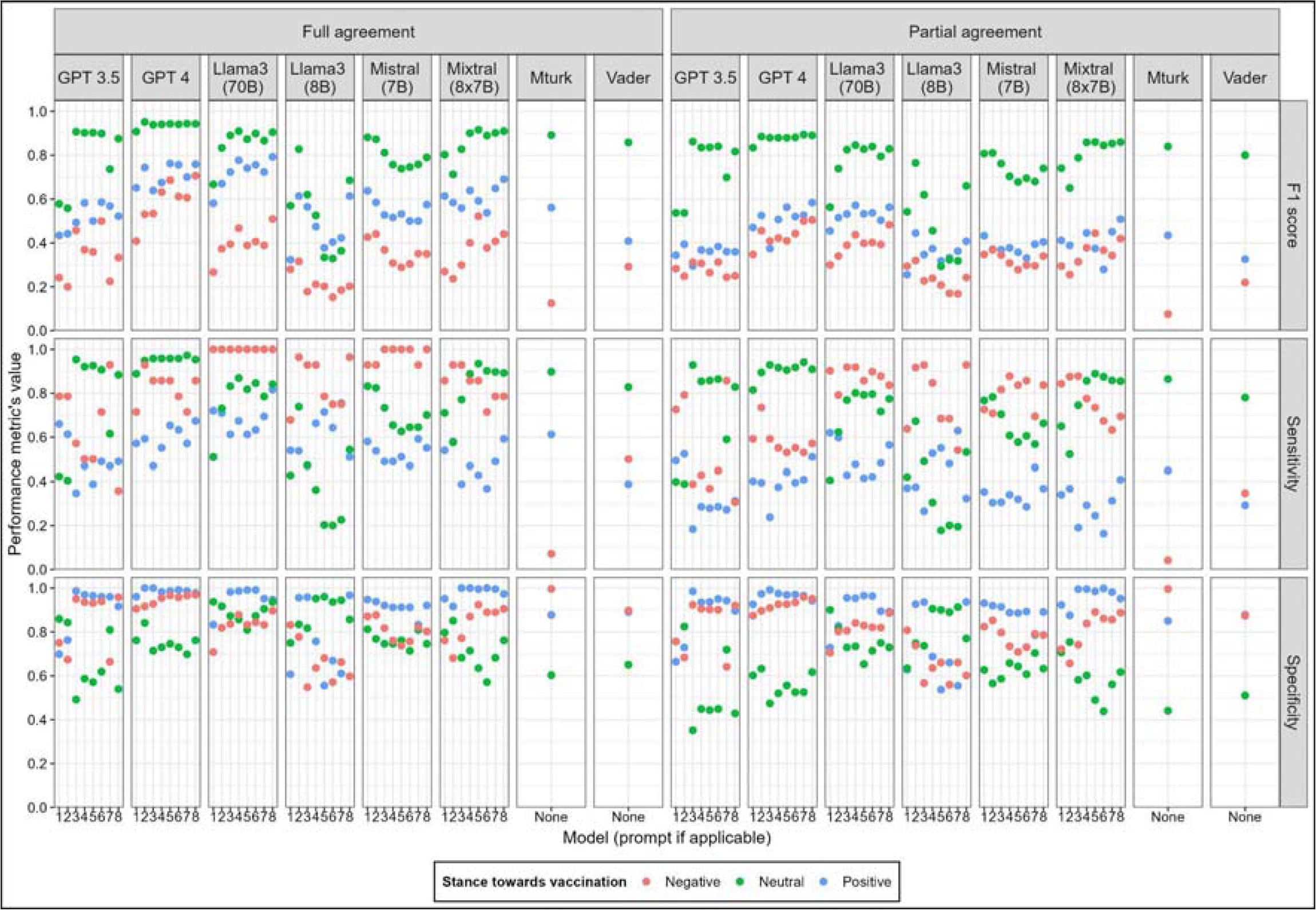
F1 score, sensitivity and specificity per model, prompt (if applicable), stance and experts’ agreement.

The performance of crowd annotators was the lowest for the minority class (negative stance towards vaccination) having the lowest F1 score and sensitivity (0.08) and the highest specificity (0.99). The metrics of the other methods and prompts had more variability when comparing among all of them. Overall, the F1 score was higher for the negative class and lower for the positive class for all methods and prompts. The sensitivity had higher variability among methods and prompts within methods with an overall tendency of higher values for neutral and negative classes. The specificity had an overall higher value for the neutral and negative classes, except for Llama3 which had a higher value for neutral and positive classes..

## Discussion

Understanding the population’s perception of public health interventions, such as vaccination, and current public health events is critical for public health experts and epidemiologists. This understanding helps them adjust risk communication and community engagement activities. Achieving this is challenging and requires a holistic approach. Our study concentrates on using social media to assess users’ stances towards vaccination. However, our method could be easily extended to cover other public health interventions and objectives, as well as other platforms or sources of information. With the continuous advancement in LLM technology and better understanding of their performance in different studies and contexts, their usage and integration into public health surveillance systems could greatly improve our ability to monitor and respond to changing public health attitudes.

Our study provides an analysis of attitudes towards vaccination, derived from 1,000 randomly selected English-language tweets collected between 2019 and 2022. This analysis is based on a consensus among four experts regarding the vaccination stance expressed in each tweet. Moreover, we extracted vaccination stances using eight alternative methods: crowd annotators, a rule-based sentiment analysis tool, and six large language models (LLMs), specifically focusing on tweets where at least three out of four experts agreed on the tweet’s classification (i.e., partial expert agreement). These eight methods were chosen because each offers unique advantages. Crowd annotators are considered the gold standard for scalable human annotation, but are nonetheless slow and costly. Rule-based sentiment analysis, while not incorporating the latest machine learning technologies, is valued for its simplicity, speed, and the ability to automate and run on local computers.

The six LLMs, which all offer automation and speed, were selected because they represent the leading edge in their field. The two GPT models from OpenAI, known for underpinning ChatGPT, are among the most recognized and utilised. GPT-3.5, the foundation of the free version of ChatGPT, and GPT-4, the most advanced model publicly available via the OpenAI platform and the basis for ChatGPT Plus, are considered state-of-the-art in LLM technology as of this writing. Access to both models is restricted to OpenAI’s platform. Mistral (7B) and Mixtral (8×7B), from Mistral AI, are considered state-of-the-art for open models. The Mistral 7B model, smaller with 7 billion parameters, can operate on a personal computer, such as a laptop. Mixtral (8×7B), a more complex “mixture of experts” model, may not run on most personal computers but can be installed on local server environments. This versatility implies that Mistral AI’s open models provide privacy benefits, as they allow data to stay on-premises. Furthermore, the two new models launched in April 2024 by Meta (Llama3 8B and 70B) are considered, at the time of writing, the state-of-the-art for open models with similar technical requirements as Mistral (7B) and Mixtral (8×7B), respectively.

We included a lexicon and rule-based sentiment analysis tool, specifically designed to extract only the general sentiment of a text, as a baseline for comparison with the more advanced LLMs. Our findings indicated that this method achieved higher accuracy than some LLMs, particularly those utilising zero-shot prompting. This suggests that inadequate prompt engineering can result in poorer outcomes than simply determining the general sentiment with a straightforward tool like VADER. Similar to crowdsourcing, VADER demonstrated high specificity but low sensitivity for minority classes, indicating that while these methods are less effective in identifying minority classes, they benefit from a reduced rate of false positive detections.

We observed a notable class imbalance among the three identified stances or classes (negative, neutral and positive), similar to what has been found in previous studies [35,36]. The majority were neutral, followed by positive, with negative being the least common stance. A high proportion of tweets (76% of the 1,000 annotated tweets) achieved a partial agreement among experts, but full agreement was limited to 37% of these. This pattern was consistent across all the other methods, with the exception of one GPT-3.5 and several Llama3 (8B) prompts showing a more balanced class distribution.

In the analysis of the tweets classified using crowdsourcing, 86% of the tweets showed partial agreement among annotators, similar to the experts pattern. However, a significantly higher 91% of these reached full agreement. This might be due to the homogeneity of Amazon Mturk annotators. There was no partial or full agreement in negative tweets, with all neutrally-classified tweets reaching full agreement. This could be attributed to our methodological choice of not including a category for unclear or irrelevant tweets as in other studies [13,14], and instructing annotators to opt for neutrality in cases of uncertainty.

In the analysis of the tweets classified using LLMs, the predominant prediction was neutral, except in GPT-3.5 zero-shot prompts where a more even class distribution was found, which could be a consequence of the simplicity of these prompts. GPT-4 demonstrated a consistent class distribution across prompts, as did Mistral (7B) and Mixtral (8×7B) for few-shot prompts, suggesting less prompt-dependency. This is a notable improvement over GPT-3.5, where simpler prompts showed a significantly different class distribution compared to more complex ones.

Some LLMs have shown similar results using different prompts, where having more examples or instructions slightly increased the performance. This is the case for GPT, Mixtral (8×7B) and Llama3 (70B). However, Mistral (7B) and Llama3 (8B), not only showed a higher prompt-dependency for their performance but also a worse performance with higher number of examples and instructions. This could be explained by being more sensitive to overfitting, in which the model relies on the type of information included in the examples focusing their learning only on these, which translates in a much worse performance when being exposed to different texts not included in the list of examples. These aspects are expected to be seen also when using LLMs for other tasks, so the results found in this study in terms of prompt-dependency and sensitivity to overfitting could be easily transferred to other studies.

All methods showed lower performance on the tweets with partial experts’ agreement, indicating a common challenge in resolving ambiguity or unclarity. In general, this ambiguity is caused by having single words with multiple meanings, i.e., polysemy, which is more noticeable in short texts as social media posts in which there is not much information on the context [37]. Other factors, such as humour, sarcasm, or general misunderstandings, indicate that a single objective assessment can be challenging, depending as much on the reader’s interpretation as on the content itself. These considerations are important when employing scalable methods and evaluating the potential risks of misclassification.

Furthermore, despite the 2-2 agreement tweets having lower overall accuracy than the partial experts’ agreement tweets, the relative comparison of accuracy among methods and prompts was very similar. This shows a stability on the expected accuracy when using a specific method and prompt, when applicable, for the same tasks, even if the agreement among experts varies.

Generally, the highest performing prompts of GPT-3.5 and GPT-4 surpassed crowdsourcing in the overall accuracy, as supported by previous studies [38–41]. Furthermore, the overall accuracy of the best performing prompts of Mixtral (8×7B) and specific accuracy for the minority class of the best performing prompt of Mistral (7B) also outperformed crowdsourcing. The cost benefit and time effectiveness of LLMs over crowd annotators, despite usage fees, has been previously highlighted [38]. Furthermore, a study has shown that up to 46% of crowd workers resorted to LLMs for task completion [42].

Each method showed varying performance per class. Some prompts of GPT-4 and Mixtral (8×7B) stood out with the highest overall accuracy and statistically significant better performance than just selecting the majority class for all tweets. They also had the highest F1 scores across all classes, indicating a reduced impact from the class imbalance. GPT-3.5 had the greatest variability in performance by class, particularly in the tweets with full agreement, showing higher dependency on the prompt than other LLMs. Crowdsourcing showed its weakest performance in the negative class, being the method with the lowest performance, suggesting a need for more diverse examples in the dataset. These differences in performance per class must be considered to use complementary models depending on their performance and the objective of the annotation as suggested in a previous study [40].

Mixtral (8×7B), an open-source “mixture of experts” model, demonstrated a performance comparable to GPT-4, a strong result given its accessibility and lower computational requirements. It showed consistent performance across different prompts, with simpler prompts often yielding better results. The accuracy of Mixtral (8×7B) in tweets with full experts’ agreement was statistically better than merely selecting the majority class, a trend also observed for many GPT-4 prompts, and some GPT 3.5 prompts. Given the open nature of Mixtral (8×7B) and its ability to be installed in a local computing environment, this makes Mixtral (8×7B) a very interesting model for use in data-sensitive situations.

Llama3 was expected to have a similar performance as Mistral (7B) and Mixtral (8×7B) based on the preliminary assessments and benchmarking of these models for other tasks [43]. On the one hand, our findings show that Llama3 (8B) had a lower performance than the other models, except for prompt 2, but had a similar relative performance among prompts as Mistral (7B), in which zero-shot prompts had better results than the few-shot prompts. On the other hand, Llama3 (70B) showed a similar performance than Mixtral (8×7B) with the same computing requirements allowing the model to be installed locally for data-sensitive situations,

The limitations of this study are that we did not analyse the explanations provided by LLMs, nor collected these from crowd annotators, which could offer insights into the discrepancies with expert classification. A study by Huang et al., 2023, suggests that GPT-3.5 provides statistically clearer explanations than human-written ones [39], indicating a potential area for future research. Additionally, while our study was limited to English tweets, a previous research indicates that GPT’s performance is consistent in non-English languages when prompts remain in English [40], allowing for multilingual studies.

Future studies could investigate the accuracy of using different methods, including LLMs with different prompts, for other public health interventions or using data from other social media platforms. Furthermore, the impact on the accuracy when using non-English tweets and/or using prompts in a different language than the tweets could be investigated. In general, the speed of the emergence of new models requires a continuous assessment of the models for a public health context.

In summary, GPT-4, Llama3 (70B) and Mixtral (8×7B) showed a consistent high performance across prompts, while GPT3.5, Llama3 (8B) and Mistral (7B) showed more variability and prompt dependency, showing a higher relevance of the prompt engineering for these models. In general, LLMs, particularly in the minority negative class, outperformed crowdsourcing, highlighting their efficacy even with limited class-specific data. Overall, the use of best-in-class models like the open Mixtral (8×7B) and Llama3 (70B) models or the closed GPT-4 model with few-shot prompting are the best methods. All current alternatives have significant risks of substantial misclassification.

## Data Availability

The data (list of tweets identifiers and summarised anonymised datasets) and R and Python code used can be found in the online repository at https://github.com/digitalepidemiologylab/llm_crowd_experts_annotation.

## Acknowledgements

We would like to thank the crowd and experts for their valuable inputs in the annotation of the tweets.

