## Supplementary material for "Use of large language models as a scalable approach to understanding public health discourse": S1 File

### **S1 Prompts used for large language models**

**Prompt 1:**

Give me the sentiment regarding vaccination in tweet text delimited by triple backticks. \

Use one of the following words: neutral, negative, positive. \

```{TWEET_TEXT_HERE}```

**Prompt 2:**

Give me the sentiment regarding vaccination expressed by the user in the tweet text delimited by triple backticks. \

Use one of the following words: neutral, negative, positive. Include an explanation for the selection of the sentiment. \

```{TWEET_TEXT_HERE}```

**Prompt 3:**

Give me the sentiment regarding vaccination expressed by the user in the tweet text delimited by triple backticks. \

Use one of the following words: neutral, negative, positive. Include an explanation for the selection of the sentiment. \

\

News, factual or objective tweets as well as generally ambiguous examples should be classified as neutral.\

\

Examples of positive tweets: \

* Some news of hope. Hopefully this vaccine becomes success and India becomes leader again. \

* This has been a horrendous year for so many people who have lost loved ones. It would be wonderful if the #vaccine was effective and could be administered world-wide to prevent further loss. My fingers and toes are crossed. \

\

Examples of neutral tweets: \

* AstraZeneca Pauses Vaccine Trial After Test Subject Gets Ill <url> \

* The federal government outlined a sweeping plan Wednesday to make vaccines for COVID-19 available for free to all Americans. <url> \

\

Examples of negative tweets: \

* There's a reason these vaccines can take up to 20 years to reach market. Say no to the vaccine. \

* Vaccines may not be good for kids like the Dengue Fever vaccine. You’re playing with fire. <url> \

\

```{TWEET_TEXT_HERE}```

**Prompt 4:**

Give me the sentiment regarding vaccination expressed by the user in the tweet text delimited by triple backticks. \

Use one of the following words: neutral, negative, positive. Include an explanation for the selection of the sentiment. \

\

News, factual or objective tweets as well as generally ambiguous examples should be classified as neutral except if:\

* User agrees with a positive statement on vaccines or vaccinations which should be classified as positive \

* User disagrees with a positive statement on vaccines or vaccinations which should be classified as negative \

* User is angry or not happy when vaccines are not available which should be classified as positive \

* User is angry or not happy with a negative statement on vaccines or vaccinations which should be classified as positive \

* User is happy with a negative statement on vaccines or vaccinations which should be classified as negative \

* User is enquiring for availability or increased availability of a vaccine which should be classified as positive \

\

Examples of positive tweets: \

* Some news of hope. Hopefully this vaccine becomes success and India becomes leader again. \

* This has been a horrendous year for so many people who have lost loved ones. It would be wonderful if the #vaccine was effective and could be administered world-wide to prevent further loss. My fingers and toes are crossed. \

* A healthy recovery of the world from COVID-19 is only possible when no one is left behind making it necessary that the vaccine is accessible to all without any hurdle. #HealthForAll #Equality #Equity #publichealth #awarness. \

\

Examples of neutral tweets: \

* AstraZeneca Pauses Vaccine Trial After Test Subject Gets Ill <url> \

* The federal government outlined a sweeping plan Wednesday to make vaccines for COVID-19 available for free to all Americans. <url> \

\

Examples of negative tweets: \

* There's a reason these vaccines can take up to 20 years to reach market. Say no to the vaccine. \

* Vaccines may not be good for kids like the Dengue Fever vaccine. You’re playing with fire. <url> \

\

```{TWEET_TEXT_HERE}```

**Prompt 5:**

Give me the sentiment regarding vaccination expressed by the user in the tweet text delimited by triple backticks. \

Use one of the following words: neutral, negative, positive. Include an explanation for the selection of the sentiment. \

\

News, factual or objective tweets as well as generally ambiguous examples should be classified as neutral except if:\

* User agrees with a positive statement on vaccines or vaccinations which should be classified as positive \

* User disagrees with a positive statement on vaccines or vaccinations which should be classified as negative \

* User is angry or not happy when vaccines are not available which should be classified as positive \

* User is angry or not happy with a negative statement on vaccines or vaccinations which should be classified as positive \

* User is happy with a negative statement on vaccines or vaccinations which should be classified as negative \

* User is enquiring for availability or increased availability of a vaccine which should be classified as positive \

\

Classify as positive if:\

* The user expresses hope or anticipation towards a vaccine.\

* The user expresses desire for having more vaccine doses. \

* The user is ironic on people being negative towards vaccine (e.g., I am not getting a vaccine because they are microchipping you. Posted on social media , on a phone..that has all your info, your location...and the info is being shared and sold ...Ummmm ok) \

\

Examples of positive tweets: \

* Some news of hope. Hopefully this vaccine becomes success and India becomes leader again. \

* This has been a horrendous year for so many people who have lost loved ones. It would be wonderful if the #vaccine was effective and could be administered world-wide to prevent further loss. My fingers and toes are crossed. \

* A healthy recovery of the world from COVID-19 is only possible when no one is left behind making it necessary that the vaccine is accessible to all without any hurdle. #HealthForAll #Equality #Equity #publichealth #awarness. \

* IF I SEE ONE MORE FUCKING POLITICIAN GET THE VACCINE BEFORE FRONT LINE WORKERS I'M GONNA LOSE MY SHIT!! \

* When will we learn to respect our scientific community ? Pathetic that ppl want to play politics on the vaccine \

* @user @user @user etc if U have so much problem so plz don't get them vaccinated, we know, U have enough money for treatment. U guys fall mild sick, immediately rush to medanta hospital.\

* $MRNA recieved approval across the pond in the U.K. demand increasing globally for a vaccine as cases rise. #stocks #finance #markets #investing #money #wealth #economy #OptionsTrading \

* On the other hand, they're more likely to receive much more effective and safe vaccines, so it's not quite as bad as you might think \

\

Examples of neutral tweets: \

* AstraZeneca Pauses Vaccine Trial After Test Subject Gets Ill <url> \

* The federal government outlined a sweeping plan Wednesday to make vaccines for COVID-19 available for free to all Americans. <url> \

\

Examples of negative tweets: \

* There's a reason these vaccines can take up to 20 years to reach market. Say no to the vaccine. \

* Vaccines may not be good for kids like the Dengue Fever vaccine. You’re playing with fire. <url> \

\

```{TWEET_TEXT_HERE}```

**Prompt 6:**

Give me the sentiment regarding vaccination expressed by the user in the tweet text delimited by triple backticks. \

Use one of the following words: neutral, negative, positive. Include an explanation for the selection of the sentiment. \

\

News, factual or objective tweets as well as generally ambiguous examples should be classified as neutral except if:\

* User agrees with a positive statement on vaccines or vaccinations which should be classified as positive \

* User disagrees with a positive statement on vaccines or vaccinations which should be classified as negative \

* User is angry or not happy when vaccines are not available which should be classified as positive \

* User is angry or not happy with a negative statement on vaccines or vaccinations which should be classified as positive \

* User is happy with a negative statement on vaccines or vaccinations which should be classified as negative \

* User is enquiring for availability or increased availability of a vaccine which should be classified as positive \

\

Classify as positive if:\

* The user expresses hope or anticipation towards a vaccine.\

* The user expresses desire for having more vaccine doses. \

\

Examples of positive tweets: \

* Some news of hope. Hopefully this vaccine becomes success and India becomes leader again. \

* This has been a horrendous year for so many people who have lost loved ones. It would be wonderful if the #vaccine was effective and could be administered world-wide to prevent further loss. My fingers and toes are crossed. \

* A healthy recovery of the world from COVID-19 is only possible when no one is left behind making it necessary that the vaccine is accessible to all without any hurdle. #HealthForAll #Equality #Equity #publichealth #awarness. \

* IF I SEE ONE MORE FUCKING POLITICIAN GET THE VACCINE BEFORE FRONT LINE WORKERS I'M GONNA LOSE MY SHIT!! \

* When will we learn to respect our scientific community ? Pathetic that ppl want to play politics on the vaccine \

* @user @user @user etc if U have so much problem so plz don't get them vaccinated, we know, U have enough money for treatment. U guys fall mild sick, immediately rush to medanta hospital.\

* $MRNA recieved approval across the pond in the U.K. demand increasing globally for a vaccine as cases rise. #stocks #finance #markets #investing #money #wealth #economy #OptionsTrading \

* On the other hand, they're more likely to receive much more effective and safe vaccines, so it's not quite as bad as you might think \

\

Examples of neutral tweets: \

* AstraZeneca Pauses Vaccine Trial After Test Subject Gets Ill <url> \

* The federal government outlined a sweeping plan Wednesday to make vaccines for COVID-19 available for free to all Americans. <url> \

\

Examples of negative tweets: \

* There's a reason these vaccines can take up to 20 years to reach market. Say no to the vaccine. \

* Vaccines may not be good for kids like the Dengue Fever vaccine. You’re playing with fire. <url> \

\

```{TWEET_TEXT_HERE}```

**Prompt 7:**

Give me the sentiment regarding vaccination expressed by the user in the tweet text delimited by triple backticks. \

Use one of the following words: neutral, negative, positive. Include an explanation for the selection of the sentiment. \

\

News, factual or objective tweets as well as generally ambiguous examples should be classified as neutral except if:\

* User agrees with a positive statement on vaccines or vaccinations which should be classified as positive \

* User disagrees with a positive statement on vaccines or vaccinations which should be classified as negative \

* User is angry or not happy when vaccines are not available which should be classified as positive \

* User is angry or not happy with a negative statement on vaccines or vaccinations which should be classified as positive \

* User is happy with a negative statement on vaccines or vaccinations which should be classified as negative \

* User is enquiring for availability or increased availability of a vaccine which should be classified as positive \

\

Classify as positive if:\

* The user expresses hope or anticipation towards a vaccine.\

* The user expresses desire for having more vaccine doses. \

\

Examples of positive tweets: \

* Some news of hope. Hopefully this vaccine becomes success and India becomes leader again. \

* This has been a horrendous year for so many people who have lost loved ones. It would be wonderful if the #vaccine was effective and could be administered world-wide to prevent further loss. My fingers and toes are crossed. \

* A healthy recovery of the world from COVID-19 is only possible when no one is left behind making it necessary that the vaccine is accessible to all without any hurdle. #HealthForAll #Equality #Equity #publichealth #awarness. \

* IF I SEE ONE MORE FUCKING POLITICIAN GET THE VACCINE BEFORE FRONT LINE WORKERS I'M GONNA LOSE MY SHIT!! \

* When will we learn to respect our scientific community ? Pathetic that ppl want to play politics on the vaccine \

* @user @user @user etc if U have so much problem so plz don't get them vaccinated, we know, U have enough money for treatment. U guys fall mild sick, immediately rush to medanta hospital.\

* $MRNA recieved approval across the pond in the U.K. demand increasing globally for a vaccine as cases rise. #stocks #finance #markets #investing #money #wealth #economy #OptionsTrading \

* On the other hand, they're more likely to receive much more effective and safe vaccines, so it's not quite as bad as you might think \

* Why are not more COVID-19 vaccine doses available immediately? <url> \

* America have fewer COVID-19 vaccines due to #MassMurderer #realDonaldTrumpâ€™s FAILED financial interests <url> \

\

Examples of neutral tweets: \

* AstraZeneca Pauses Vaccine Trial After Test Subject Gets Ill <url> \

* The federal government outlined a sweeping plan Wednesday to make vaccines for COVID-19 available for free to all Americans. <url> \

\

Examples of negative tweets: \

* There's a reason these vaccines can take up to 20 years to reach market. Say no to the vaccine. \

* Vaccines may not be good for kids like the Dengue Fever vaccine. You’re playing with fire. <url> \

* Dr: Are you experiencing any side effects from the vaccine? Me: * sleeps while standing there* huh? \

* There is a virus which has killed so many people for years. This virus is called hunger and the vaccine for hunger is food. However no one talks about it, why? Because this virus doesn't kill rich people.\

* @user : There are over 100 different coronavirus vaccine candidates in the works. These candidates take a variety of approaches to protecting the body against COVID-19: <url> #CoronaVirusUpdates #CoronavirusVaccine #StayHome <url> \

* <It would go away without the vaccine> #HerdMentality this man knows nothing about viruses. \

* Anti-vaxxer Novak Djokovic. #COVID19 #tennis The current world number one is the fourth player to test positive from the event that did not even pay lip service to social distancing. <url> via @user \

\

```{TWEET_TEXT_HERE}```

**Prompt 8:**

Give me the stance of the user towards vaccination in the tweet text delimited by triple backticks, regardless of the general sentiment expressed in the tweet. A tweet with a negative general sentiment can express a positive stance of the user towards vaccination and viceversa. \

Use one of the following words: neutral, negative, positive. Include an explanation for the selection of the stance. \

The reply must contain the selected word for the stance followed by '.' and followed by the explanation for the selected stance. \

\

News, factual or objective tweets as well as generally ambiguous examples should be classified as neutral except if:\

* User agrees with a positive statement on vaccines or vaccinations which should be classified as positive \

* User disagrees with a positive statement on vaccines or vaccinations which should be classified as negative \

* User is angry or not happy when vaccines are not available which should be classified as positive \

* User is angry or not happy with a negative statement on vaccines or vaccinations which should be classified as positive \

* User is happy with a negative statement on vaccines or vaccinations which should be classified as negative \

* User is enquiring for availability or increased availability of a vaccine which should be classified as positive \

\

Classify as positive if:\

* The user expresses hope or anticipation towards a vaccine.\

* The user expresses desire for having more vaccine doses. \

* The user is ironic on people being negative towards vaccine (e.g., I am not getting a vaccine because they are microchipping you. Posted on social media , on a phone..that has all your info, your location...and the info is being shared and sold ...Ummmm ok) \

\

Examples of positive tweets: \

* Some news of hope. Hopefully this vaccine becomes success and India becomes leader again. \

* This has been a horrendous year for so many people who have lost loved ones. It would be wonderful if the #vaccine was effective and could be administered world-wide to prevent further loss. My fingers and toes are crossed. \

* A healthy recovery of the world from COVID-19 is only possible when no one is left behind making it necessary that the vaccine is accessible to all without any hurdle. #HealthForAll #Equality #Equity #publichealth #awarness. \

* IF I SEE ONE MORE FUCKING POLITICIAN GET THE VACCINE BEFORE FRONT LINE WORKERS I'M GONNA LOSE MY SHIT!! \

* When will we learn to respect our scientific community ? Pathetic that ppl want to play politics on the vaccine \

* @user @user @user etc if U have so much problem so plz don't get them vaccinated, we know, U have enough money for treatment. U guys fall mild sick, immediately rush to medanta hospital.\

* $MRNA recieved approval across the pond in the U.K. demand increasing globally for a vaccine as cases rise. #stocks #finance #markets #investing #money #wealth #economy #OptionsTrading \

* On the other hand, they're more likely to receive much more effective and safe vaccines, so it's not quite as bad as you might think \

\

Examples of neutral tweets: \

* AstraZeneca Pauses Vaccine Trial After Test Subject Gets Ill <url> \

* The federal government outlined a sweeping plan Wednesday to make vaccines for COVID-19 available for free to all Americans. <url> \

\

Examples of negative tweets: \

* There's a reason these vaccines can take up to 20 years to reach market. Say no to the vaccine. \

* Vaccines may not be good for kids like the Dengue Fever vaccine. You’re playing with fire. <url> \

\

```{TWEET_TEXT_HERE}```
